# UGB-SegNet: Uncertainty-Guided Multi-Scale Attention and Boundary-Aware Learning for Breast Ultrasound Lesion Segmentation

**DOI:** 10.64898/2026.10.03.26364633

**Authors:** Taukir Alam, Fang-Rong Hsu

## Abstract

Accurate breast lesion segmentation in ultrasound is difficult because speckle noise, heterogeneous lesion appearance, and poorly defined margins can obscure clinically relevant boundaries. This work presents UGB-SegNet, an uncertainty-guided breast ultrasound segmentation network that combines four components: multi-scale convolutional block attention (CBAM) at three EfficientNet-B0 encoder stages, a learnable feature pyramid network for adaptive multi-scale fusion, an entropy-guided decoder that routes fine-grained encoder information toward uncertain regions, and a boundary-aware composite loss with deep supervision. The model was evaluated on the public BUSI dataset (647 annotated lesion images) using a stratified 454/64/129 train/validation/test split. On the held-out test set, UGB-SegNet achieved 81.30% Dice, 68.49% IoU, 81.71% sensitivity, 98.14% specificity, and 15.63-pixel HD95. Under identical training conditions it outperformed five directly trained baselines: U-Net, Attention U-Net, SegNet, DeepLabV3+, and TransUNet. A five-fold resampling experiment over the development set with the held-out test set fixed yielded a mean Dice of 76.36% +/-1.66% (95% CI 74.90-77.81). Reported ablations attribute gains of +2.06 Dice points to multi-scale CBAM, +1.52 to learnable FPN fusion, +1.22 to the uncertainty-guided decoder, and +1.24 to boundary-aware training. UGB-SegNet contains 14.00M parameters and runs at 62.4 FPS at 224 × 224 resolution on the tested NVIDIA GPU, providing a compact accuracy-efficiency trade-off for future prospective clinical validation.

## 1. Introduction

Breast cancer remains a major global health burden, and imaging is central to screening, diagnosis, treatment planning, and longitudinal assessment [1]. Breast ultrasound is especially useful as a complementary modality because it is non-ionising, portable, relatively inexpensive, and capable of depicting lesions in dense breast tissue. At the same time, ultrasound interpretation is affected by speckle noise, variable acquisition conditions, tissue heterogeneity, and operator dependence. These factors make consistent delineation of lesion boundaries challenging [2].

Automated segmentation can support lesion measurement, morphology assessment, computer-aided diagnosis, and subsequent classification. Deep convolutional models have therefore become a major approach to medical image segmentation [3]. U-Net and its variants established encoder-decoder architectures with skip connections as strong baselines [5], while attention gates [6], atrous spatial pyramid pooling [7], encoder-decoder index transfer [8], and CNN-Transformer hybrids [9] have expanded the design space. However, breast ultrasound remains difficult because diagnostically useful information occurs at multiple scales: fine texture and margin irregularity, intermediate lesion shape and orientation, and coarse semantic context.

This preprint introduces UGB-SegNet, a compact segmentation architecture derived from the Phase 2 work of the author’s doctoral dissertation. The design is motivated by four limitations identified in earlier breast-ultrasound segmentation work: attention restricted to a single scale, fixed multi-scale fusion, absence of uncertainty-aware refinement, and insufficient emphasis on lesion boundaries during optimisation. The proposed network addresses these limitations through multi-scale CBAM attention, learnable feature-pyramid fusion, an entropy-guided decoder, and a boundary-aware composite loss.

The main contributions are: (1) simultaneous CBAM attention at three EfficientNet-B0 encoder scales; (2) trainable fusion weights that learn the relative contribution of fine, intermediate, and coarse features; (3) attention-derived entropy maps used as an uncertainty proxy to gate decoder skip connections; (4) a boundary-aware loss that upweights lesion-margin errors by 5× and is combined with deep supervision; and (5) evaluation against five directly trained baseline architectures, five-fold development-set resampling, complexity analysis, and qualitative uncertainty analysis.

## 2. Related Work

### 2.1 Breast ultrasound segmentation

The BUSI dataset released by Al-Dhabyani et al. has become a widely used public benchmark for breast ultrasound analysis [4]. Classical encoder-decoder approaches such as U-Net [5] and SegNet [8] provide strong reference architectures. Attention U-Net introduced learned gating of skip features [6], while DeepLabV3+ combines atrous convolution and multi-scale context aggregation [7]. TransUNet uses a CNN encoder with Transformer-based global context modelling [9]. The author’s earlier Phase 1 work compared six semantic-segmentation models on a clinical dataset from Changhua Christian Hospital and identified U-Net3+ as the strongest baseline in that study [24]. The present work uses those observations as motivation but evaluates the proposed architecture primarily on BUSI.

### 2.2 Multi-scale attention, adaptive fusion, uncertainty, and boundary learning

CBAM sequentially applies channel and spatial attention and can improve feature selectivity with modest overhead [10]. Feature Pyramid Networks aggregate information across resolutions [11], and BiFPN demonstrated that learnable non-negative fusion weights can improve multi-scale feature aggregation [12]. Uncertainty estimation in deep learning is commonly approached through techniques such as Monte Carlo dropout or deep ensembles [13,14]. UGB-SegNet instead derives an entropy-like map from spatial attention and uses it as an uncertainty proxy inside the decoder. Boundary-aware losses complement region-overlap objectives by explicitly penalising contour errors [15], while focal-style objectives can reduce the dominance of easy background pixels in imbalanced segmentation [16].

## 3. Materials and Methods

### 3.1 Datasets and experimental split

The primary benchmark was the Breast Ultrasound Images (BUSI) dataset collected at the Baheya Foundation for Early Detection and Treatment of Women’s Cancer in Egypt [4]. After excluding normal examinations without a lesion mask, the dissertation used 647 annotated images: 437 benign and 210 malignant. Images were stratified with random seed 42 into 454 training images (70%), 64 validation images (10%), and 129 held-out test images (20%). Validation and test images were not augmented. The BUS-BRA dataset was used as supplementary breast-domain pre-training data for decoder initialisation [19].

**Figure 1.**
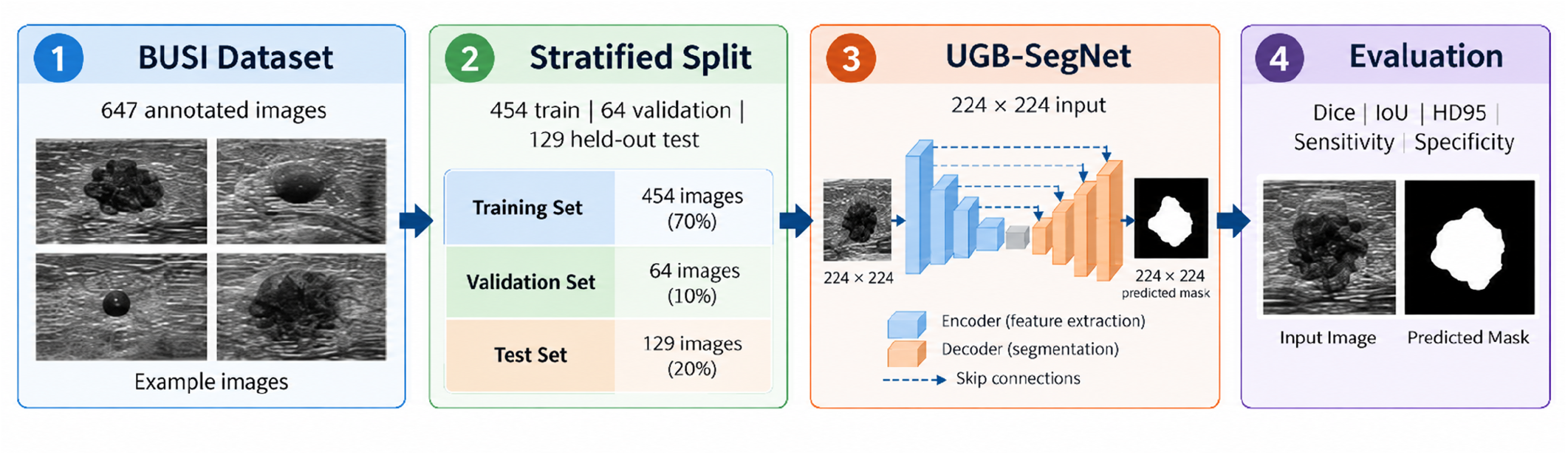
Experimental workflow for BUSI training, held-out testing, and controlled baseline evaluation.

**Table 1.**
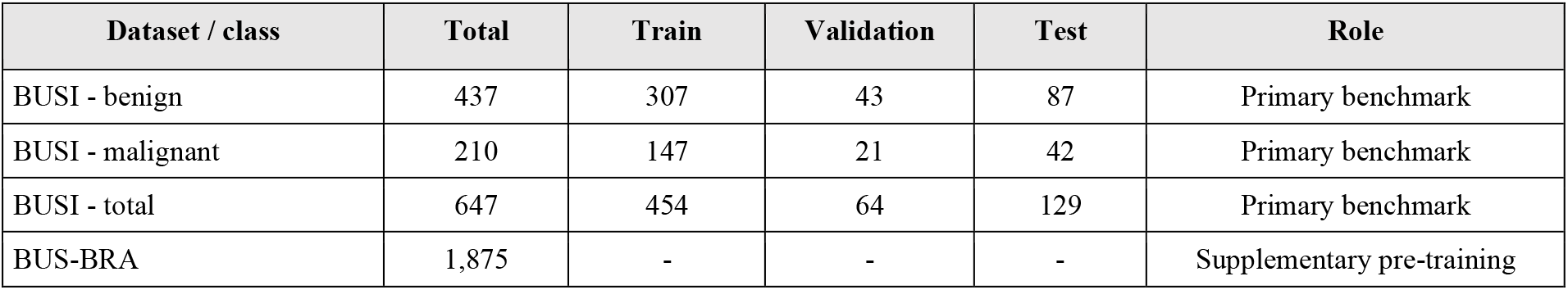
Dataset composition and BUSI experimental split.

**Figure 2.**
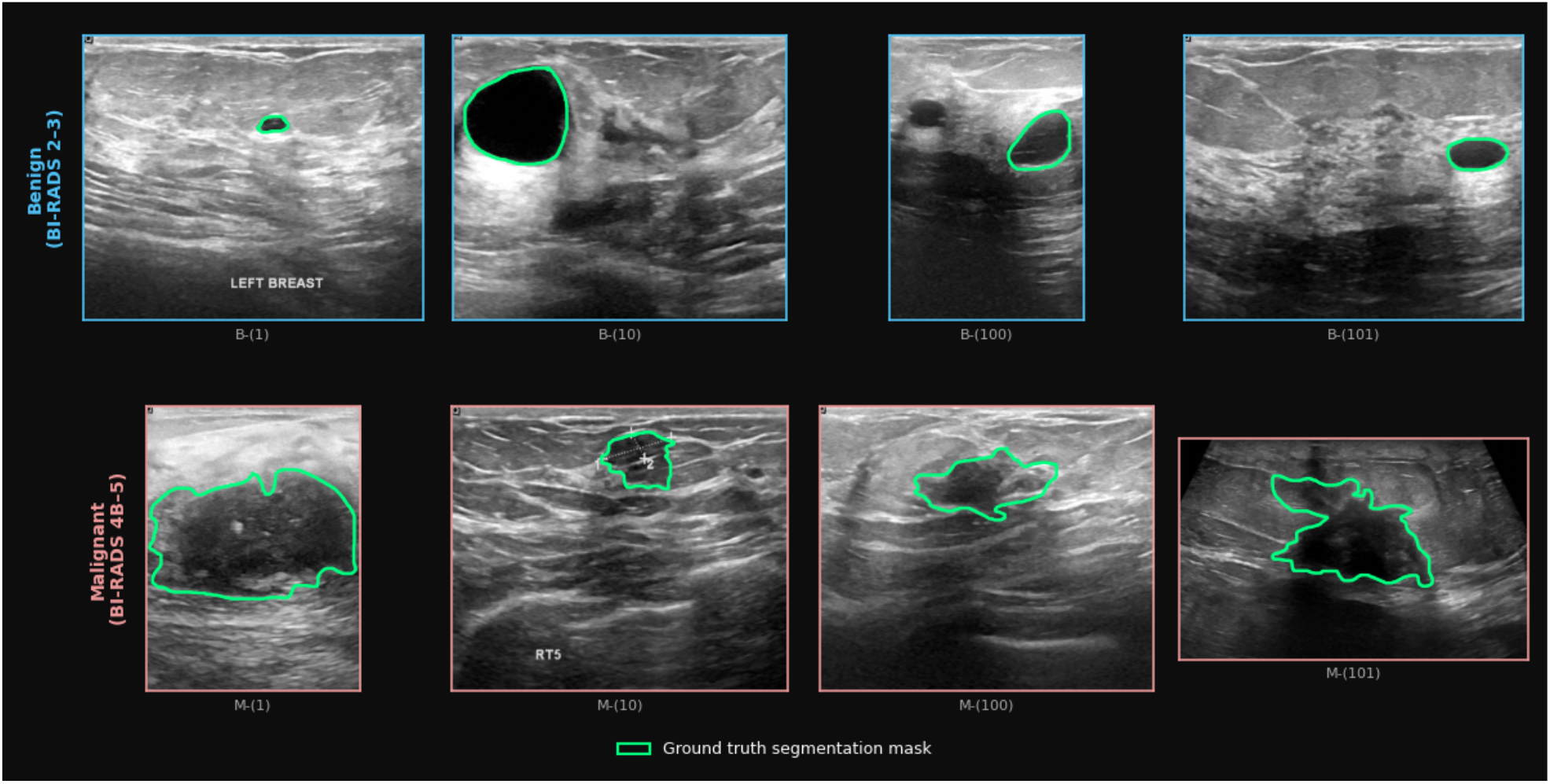
Representative BUSI benign and malignant ultrasound images with expert lesion contours.

### 3.2 Preprocessing and augmentation

Images were resized to 224×224 pixels using bilinear interpolation; binary masks were resized with nearest-neighbour interpolation to preserve label integrity. RGB channels were retained to remain compatible with ImageNet-pretrained EfficientNet-B0 weights, and inputs were normalised using ImageNet mean and standard deviation. Masks were binarised and, for images with more than one annotated lesion, lesion masks were merged by logical OR. Training-only augmentation was implemented on the fly using Albumentations [20] and included horizontal/vertical flips, random rotation (±30°), elastic deformation, brightness/contrast perturbation, Gaussian noise, and random crop/zoom.

### 3.3 UGB-SegNet architecture

UGB-SegNet uses an ImageNet-pretrained EfficientNet-B0 encoder [17]. Three intermediate feature stages are selected: s2 (24 channels, 56×56), s5 (48 channels, 28×28), and s8 (1280 channels, 7×7). These stages represent progressively larger receptive fields corresponding to fine margin texture, lesion shape and orientation, and coarse tissue semantics. The complete model contains 14.00M trainable parameters.

**Figure 3.**
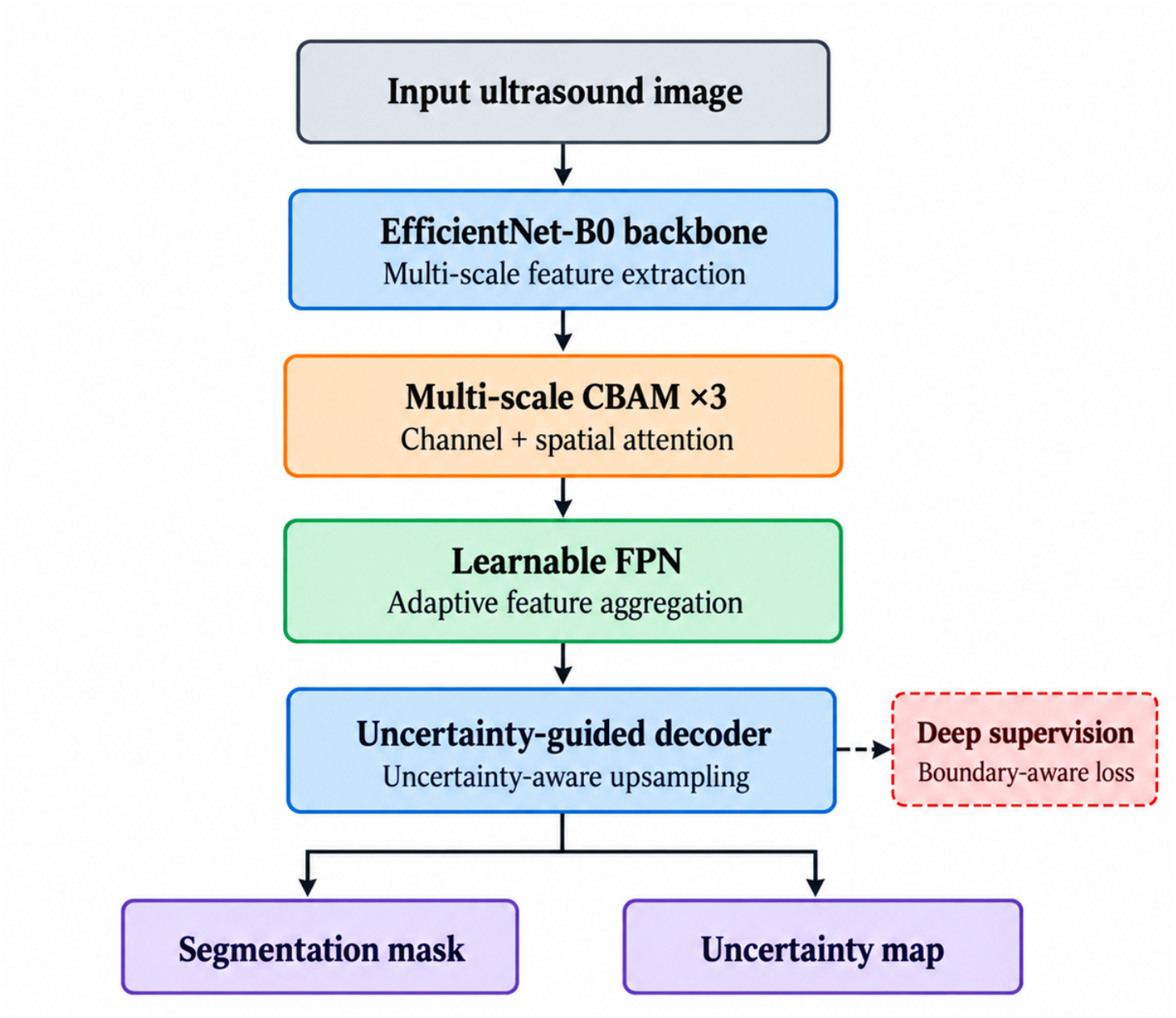
UGB-SegNet architecture. Multi-scale EfficientNet-B0 features are refined with CBAM, fused by a learnable feature pyramid, and decoded using uncertainty-guided skip routing with deep supervision and boundary-aware training.

#### 3.3.1 Multi-scale CBAM attention

A CBAM module [10] is applied independently to each of the three selected encoder stages. Channel attention emphasizes informative feature channels, whereas spatial attention emphasizes diagnostically relevant image locations. Applying attention at three scales is intended to capture fine boundary texture, intermediate morphology, and coarse semantic context simultaneously. The spatial attention output from the fine-scale stage is also transformed into an entropy-like ambiguity map; locations with intermediate attention probabilities are treated as more uncertain.

#### 3.3.2 Learnable feature-pyramid fusion

The three attended feature maps are first projected to 256 channels using 1×1 convolutions and spatially aligned to 56×56. Instead of equal-weight fusion, UGB-SegNet learns three non-negative scalar weights. The normalised learned weights reported in the dissertation were approximately 0.38, 0.35, and 0.27 for the fine, intermediate, and coarse scales, respectively. This mechanism follows the general principle of learnable multi-scale fusion used in BiFPN [12] while keeping the parameter overhead negligible.

#### 3.3.3 Uncertainty-guided decoder

The fused representation is progressively upsampled to the original 224×224 resolution. At each decoder stage, an uncertainty-derived gate modulates the corresponding encoder skip features before concatenation. In high-uncertainty regions, more fine-grained encoder detail is admitted; in lower-uncertainty regions, the decoder relies more strongly on its semantic representation. Decoder widths progressively narrow across stages, and three auxiliary prediction heads provide deep supervision during training [18]. Only the final segmentation output is used at inference.

#### 3.3.4 Boundary-aware composite loss

The training objective combines Dice loss, focal loss (alpha=0.25, gamma=2.0), a morphological boundary-weighting term, and auxiliary deep-supervision losses. A thin boundary region is generated from the target mask using dilation radius 5 and erosion radius 1; errors in this boundary region are weighted 5× relative to interior pixels. The deep-supervision contribution is weighted by 0.40. This design combines region-overlap optimisation with explicit emphasis on lesion contours [15,16].

**Figure 4.**
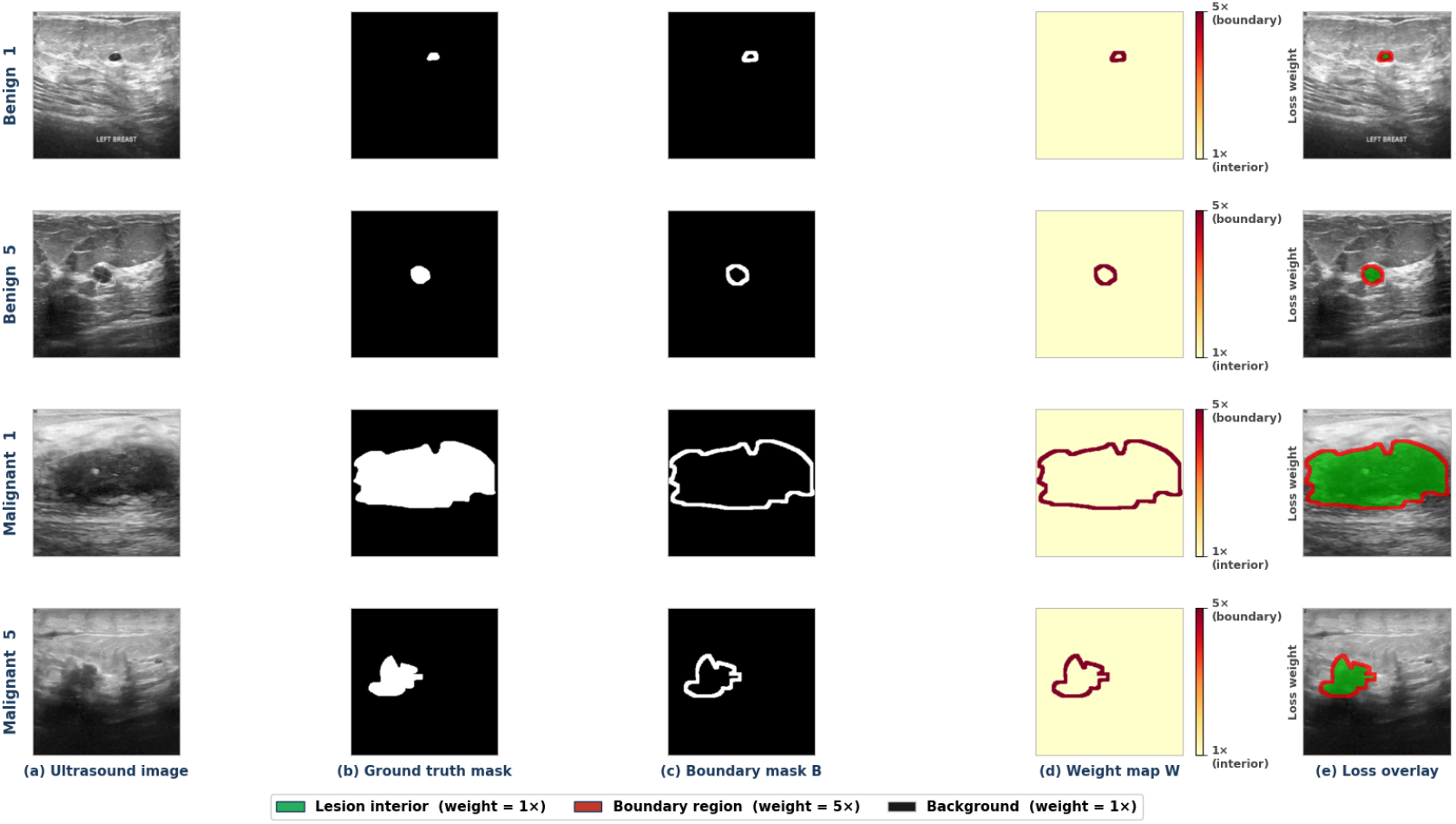
Boundary-aware loss construction. A morphological boundary ring is derived from the ground-truth mask and assigned increased training weight to emphasize lesion-margin errors.

### 3.4 Training protocol

Training used PyTorch 1.13 with CUDA 11.7 on NVIDIA RTX 3070/3080-class hardware. The batch size was 16 and optimisation used AdamW with initial learning rate 1×10^-4 and weight decay 1×10^-5. A cosine-annealing schedule reduced the learning rate toward 1×10^-6. Training was conducted for up to 80 epochs with early stopping (patience 25). During epochs 1-40, EfficientNet-B0 stages s0-s4 were frozen; during epochs 41-80, all layers were unfrozen for end-to-end fine-tuning. The best checkpoint was selected using validation Dice.

**Table 2.** Core training configuration.

| Setting | Value |
| --- | --- |
| Input resolution | 224×224×3 |
| Batch size | 16 |
| Optimiser | AdamW |
| Initial learning rate | $1 \times 10^{-4}$ |
| Weight decay | $1 \times 10^{-5}$ |
| Schedule | Cosine annealing; eta_min=$1 \times 10^{-6}$ |
| Epochs | 80 max; early stopping patience=25 |
| Frozen stage | Epochs 1-40: EfficientNet s0-s4 frozen |
| Fine-tuning | Epochs 41-80: all layers unfrozen |
| Deep-supervision weight | 0.40 |
| Boundary weight | 5× |
| Random seed | 42 |

**Figure 5.**
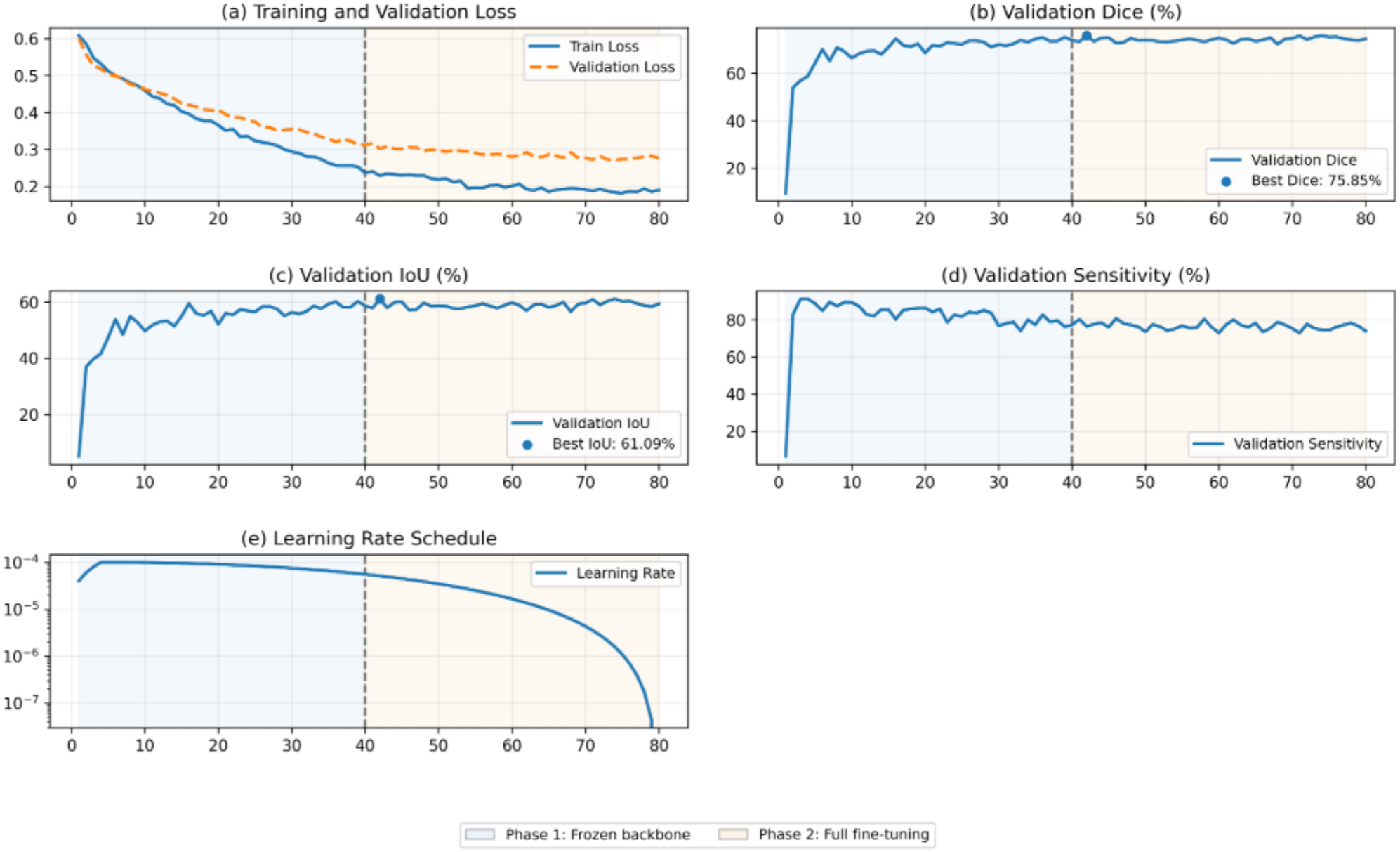
Training and validation curves for the two-phase 80-epoch schedule. The vertical dashed line indicates the transition from frozen-backbone training to full fine-tuning.

### 3.5 Evaluation and statistical analysis

Binary predictions were obtained by thresholding sigmoid outputs at 0.5. The primary overlap metric was Dice similarity coefficient, Dice = 2TP/(2TP + FP + FN). Additional metrics were intersection over union (IoU), sensitivity, specificity, and the 95th-percentile Hausdorff distance (HD95) in pixels. The dissertation reports 1,000-resample bootstrap confidence intervals and paired Wilcoxon signed-rank tests on per-image Dice scores, with alpha=0.05 [21,22]. Five published baseline architectures were trained under the same data split, optimiser, scheduler, and augmentation conditions to support a controlled architectural comparison.

**Table 3.** Directly trained baseline models.

| Model | Architecture / backbone | Parameters (M) | Key characteristic |
| --- | --- | --- | --- |
| U-Net | VGG-style encoder-decoder | 31.04 | Same-scale skip connections |
| Attention U-Net | VGG-style + attention gates | 31.04 | Attention-gated skips |
| SegNet | VGG-16 encoder-decoder | 28.81 | Pooling-index upsampling |
| DeepLabV3+ | ResNet-50 + ASPP | 39.60 | Atrous multi-scale context |
| TransUNet | ResNet50 + ViT-B/16 | 87.20 | CNN-Transformer hybrid |
| **UGB-SegNet** | **EfficientNet-B0 + proposed modules** | **14.00** | **Multi-scale attention + adaptive fusion + uncertainty guidance** |

## 4. Results

### 4.1 Held-out BUSI test performance

On the 129-image held-out BUSI test set, UGB-SegNet achieved 81.30% Dice, 68.49% IoU, 81.71% sensitivity, 98.14% specificity, and 15.63-pixel HD95. Among the five architectures trained under the same experimental protocol, DeepLabV3+ was the closest baseline in overlap performance (79.28% Dice), whereas U-Net achieved 71.24% Dice. UGB-SegNet also yielded the lowest boundary error. The dissertation reports statistically significant Dice improvements over all five baselines (p<0.01, paired Wilcoxon signed-rank test).

**Figure 6.**
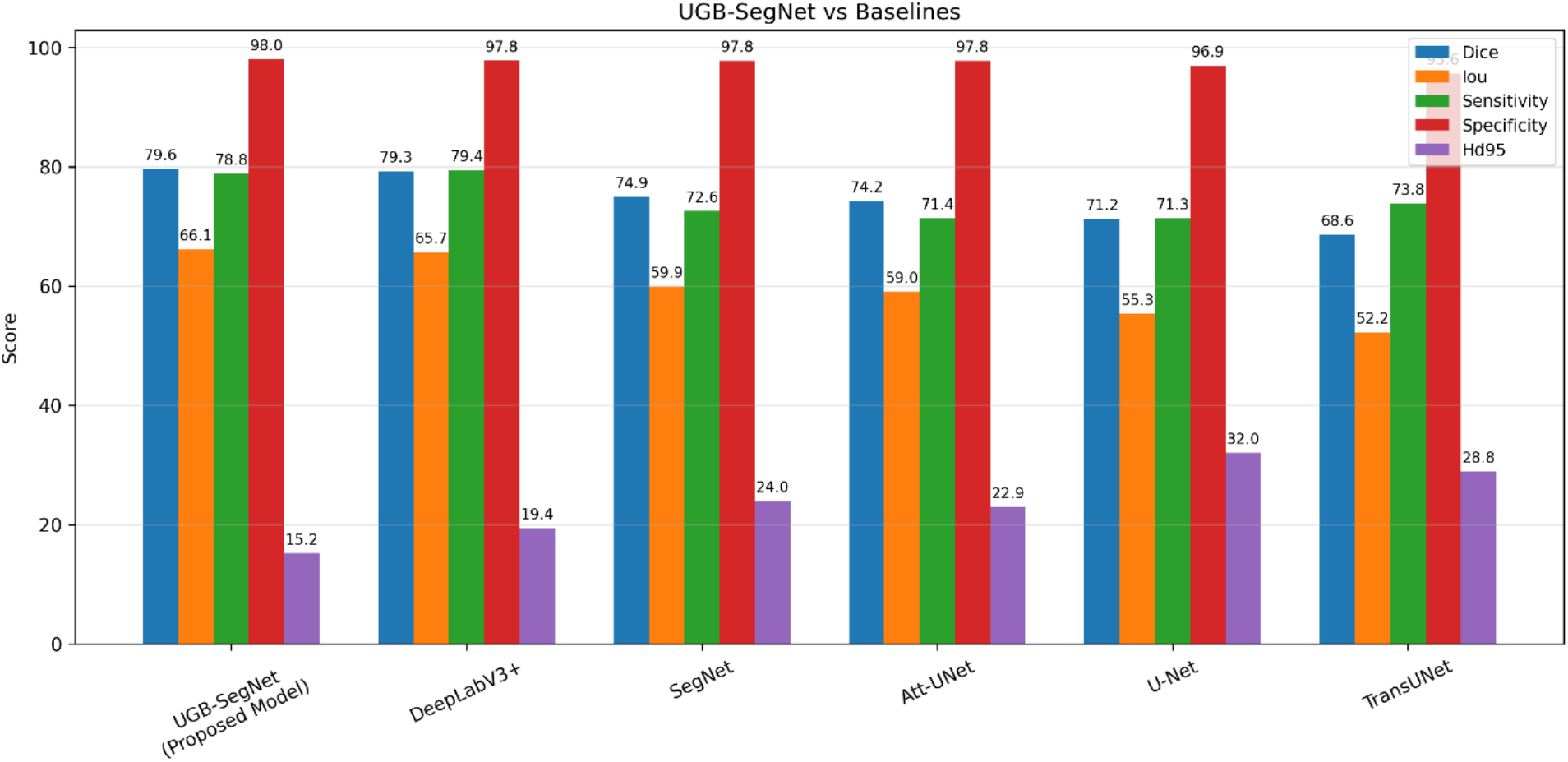
Grouped comparison of UGB-SegNet with five baseline models on the BUSI held-out test set. Dice, IoU, sensitivity, and specificity are percentages; HD95 is reported in pixels (lower is better).

**Table 4.**
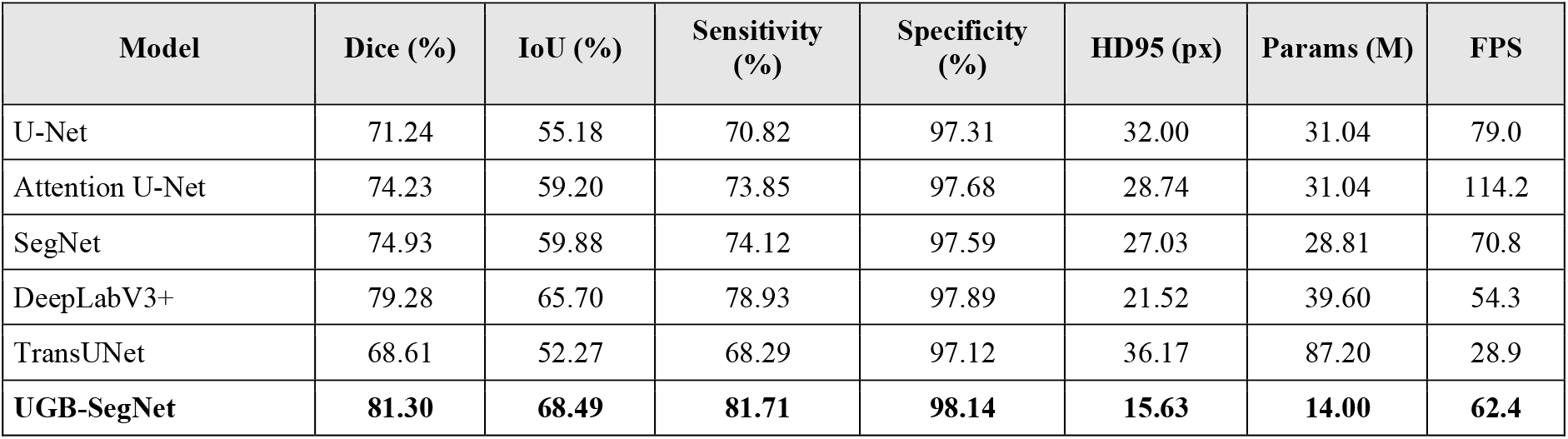
Quantitative comparison on the BUSI held-out test set (n=129).

| Model | Dice (%) | IoU (%) | Sensitivity (%) | Specificity (%) | HD95 (px) | Params (M) | FPS |
| --- | --- | --- | --- | --- | --- | --- | --- |
| U-Net | 71.24 | 55.18 | 70.82 | 97.31 | 32.00 | 31.04 | 79.0 |
| Attention U-Net | 74.23 | 59.20 | 73.85 | 97.68 | 28.74 | 31.04 | 114.2 |
| SegNet | 74.93 | 59.88 | 74.12 | 97.59 | 27.03 | 28.81 | 70.8 |
| DeepLabV3+ | 79.28 | 65.70 | 78.93 | 97.89 | 21.52 | 39.60 | 54.3 |
| TransUNet | 68.61 | 52.27 | 68.29 | 97.12 | 36.17 | 87.20 | 28.9 |
| **UGB-SegNet** | **81.30** | **68.49** | **81.71** | **98.14** | **15.63** | **14.00** | **62.4** |

### 4.2 Five-fold development-set resampling

The combined training and validation pool (n=518) was partitioned into five stratified folds, while the same 129-image held-out test set remained fixed. Across the five independently trained models, mean test Dice was 76.36% ± 1.66% with a reported 95% confidence interval of 74.90-77.81. Mean IoU was 66.83% ± 1.84%, mean sensitivity was 79.30% ± 2.30%, and mean HD95 was 24.93 ± 10.75 pixels. The spread across folds indicates sensitivity to the composition and size of the development subset, particularly for boundary distance.

**Figure 7.**
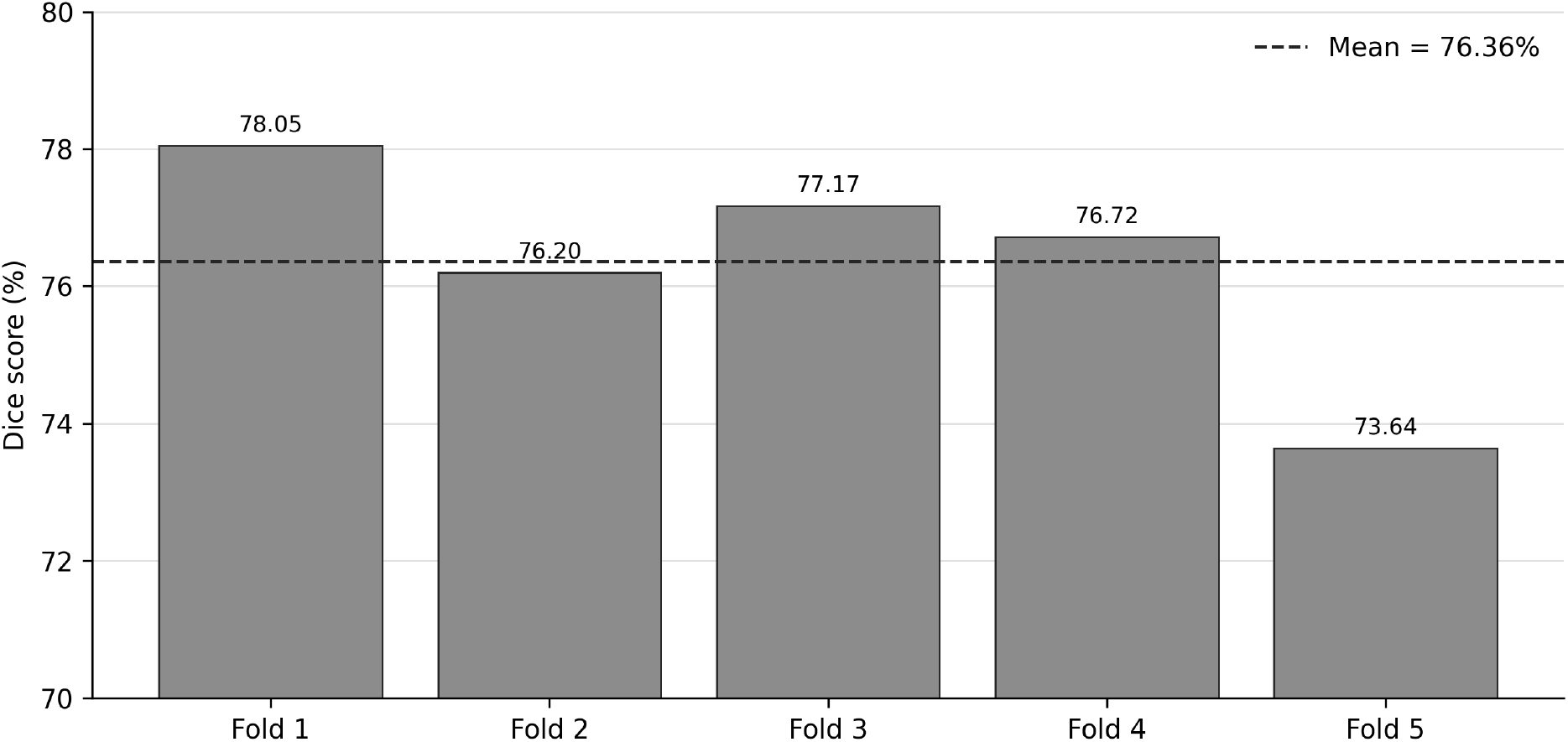
Fold-wise Dice scores from five development-set resampling runs evaluated on the fixed held-out test set. The dashed line denotes the mean Dice of 76.36%.

**Table 5.** Five-fold development-set resampling with a fixed held-out test set.

| Fold | Train | Val | Test | Dice (%) | IoU (%) | Sensitivity (%) | HD95 (px) |
| --- | --- | --- | --- | --- | --- | --- | --- |
| 1 | 414 | 104 | 129 | 78.05 | 68.93 | 77.98 | 17.75 |
| 2 | 414 | 104 | 129 | 76.20 | 66.54 | 78.61 | 22.62 |
| 3 | 414 | 103 | 129 | 77.17 | 67.76 | 82.28 | 20.05 |
| 4 | 415 | 105 | 129 | 76.72 | 66.93 | 76.62 | 20.32 |
| 5 | 415 | 105 | 129 | 73.64 | 63.97 | 81.01 | 43.91 |
| Mean ± SD | - | - | - | 76.36 ± 1.66 | 66.83 ± 1.84 | 79.30 ± 2.30 | 24.93 ± 10.75 |
| 95% CI | - | - | - | 74.90-77.81 | 65.21-68.44 | 75.28-81.32 | 15.51-34.35 |

### 4.3 Component-level effects reported in the dissertation

The dissertation reports incremental component effects from ablation experiments. Because the source document provides the component-wise deltas but not a single consolidated ablation table with all absolute intermediate configurations, the effects are reproduced here as reported deltas rather than reconstructed absolute scores.

**Figure 8.**
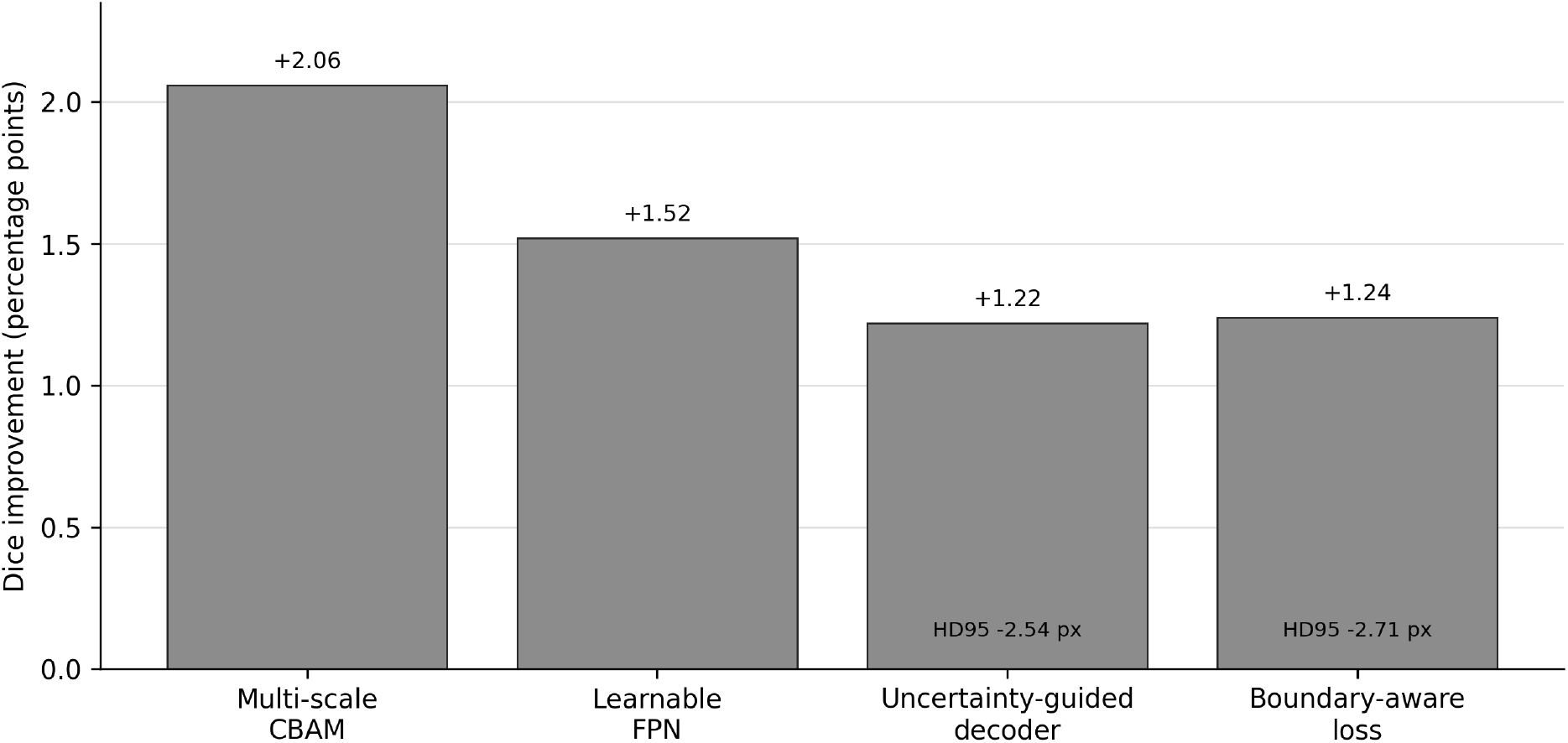
Component-level Dice improvements reported in the dissertation ablation analysis. The uncertainty-guided decoder and boundary-aware loss also reduced HD95 by 2.54 px and 2.71 px, respectively.

**Table 6.** Reported contribution of the four proposed components.

| Component | Reported effect | Primary interpretation |
| --- | --- | --- |
| Multi-scale CBAM | +2.06 Dice points | Improved multi-scale feature selection |
| Learnable FPN | +1.52 Dice points | Adaptive weighting of spatial scales |
| Uncertainty-guided decoder | +1.22 Dice; -2.54 px HD95 | Targeted refinement of ambiguous regions |
| Boundary-aware loss | +1.24 Dice; -2.71 px HD95 | Stronger contour optimisation |

### 4.4 Model complexity and runtime

UGB-SegNet contains 14.00M parameters, occupies 56.6 MB on disk, and requires 24.4 GFLOPs per 224×224 image according to the dissertation complexity analysis. Inference latency was 16.0 ms per image (62.4 FPS) on the tested NVIDIA hardware. The resulting Dice-per-parameter ratio was 5.81, substantially higher than the directly compared baselines.

**Figure 9.**
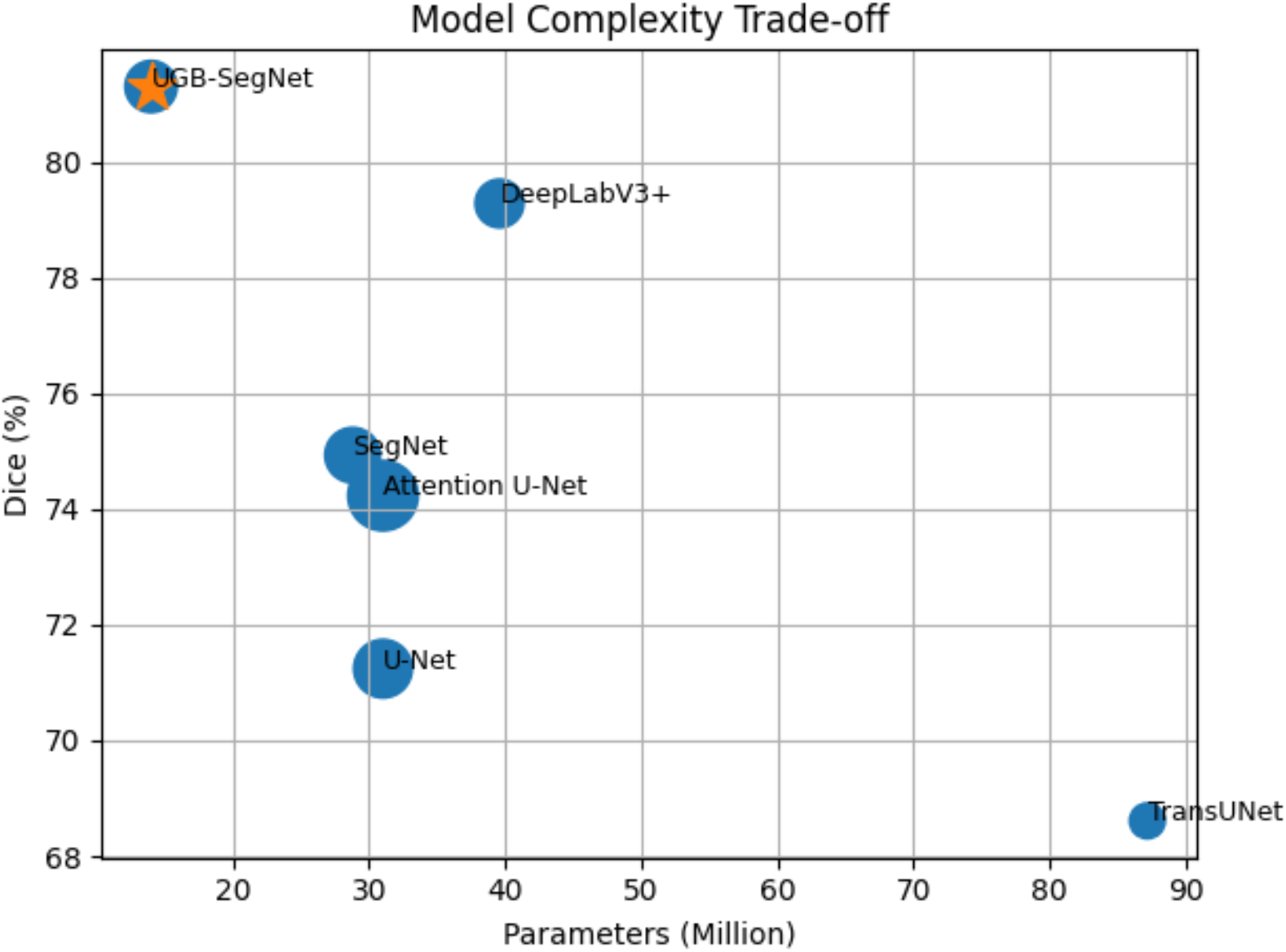
Accuracy-complexity trade-off for the directly evaluated models, showing Dice score as a function of model parameter count.

### 4.5 Uncertainty analysis

The dissertation groups predictions by segmentation quality and analyses the attention-derived entropy proxy. Mean entropy increased from approximately 0.060 in high-Dice cases to 0.219 in moderate cases, 0.388 in low-Dice cases, and 0.523 in failed cases. The inverse relationship between Dice and entropy suggests that the uncertainty proxy captures at least part of the model’s segmentation difficulty and may be useful for flagging low-confidence cases for review.

**Figure 10.**
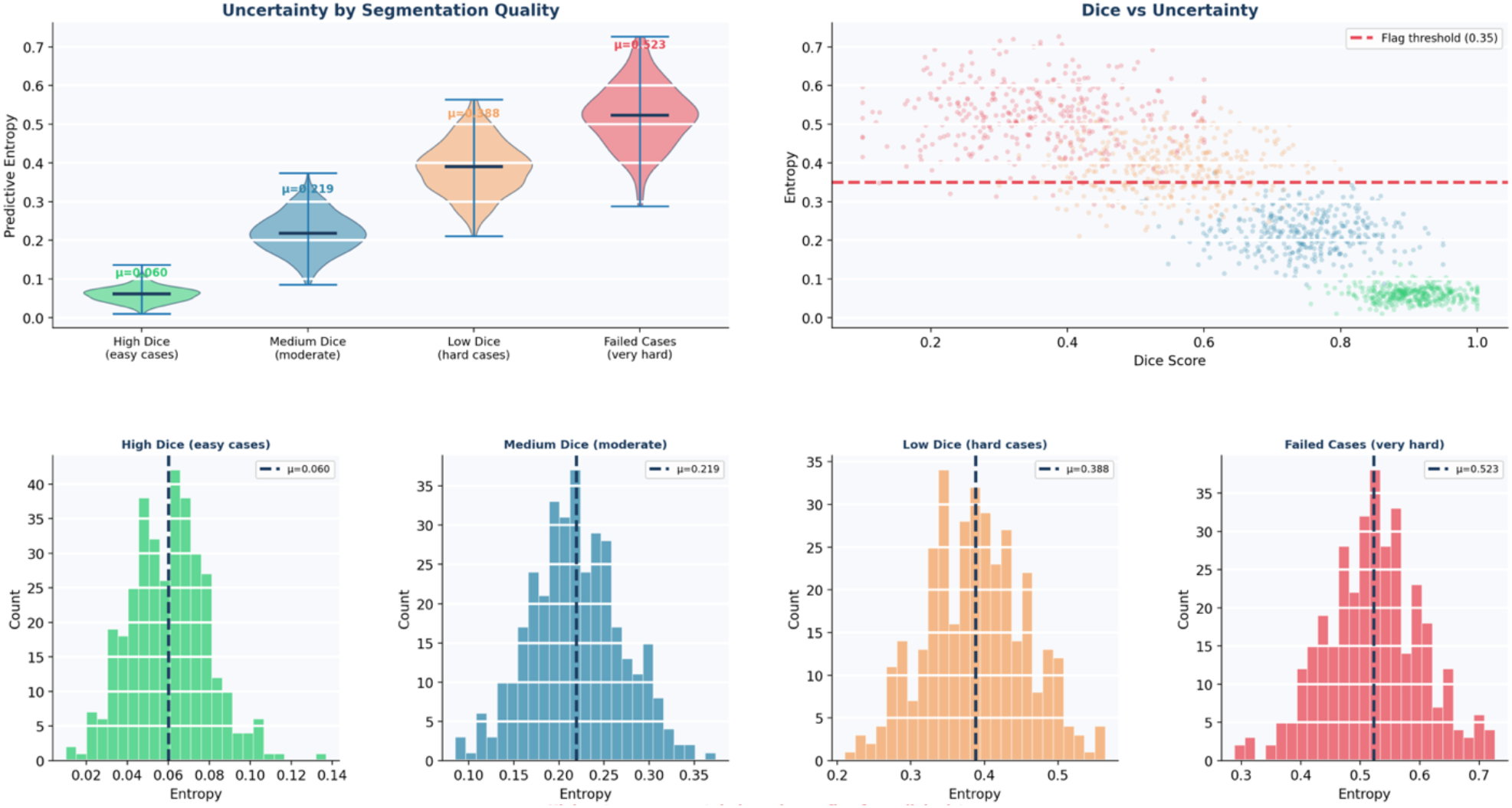
Predictive-entropy analysis across segmentation-quality groups. Higher entropy is associated with lower Dice scores and failed segmentations in the dissertation analysis.

## 5. Discussion

UGB-SegNet was designed to improve both lesion overlap and boundary precision without increasing model size. The controlled BUSI comparison supports this objective: the proposed model improved Dice by 2.02 percentage points over DeepLabV3+ and reduced HD95 from 21.52 to 15.63 pixels, while using substantially fewer parameters. Relative to U-Net, the Dice improvement was 10.06 percentage points and HD95 was reduced by 16.37 pixels. These results suggest that the combined emphasis on multi-scale selection, adaptive feature fusion, uncertainty-aware routing, and boundary-weighted optimisation is useful for breast ultrasound lesion segmentation.

The learnable fusion weights are also informative. The reported distribution (fine 0.38, intermediate 0.35, coarse 0.27) places the highest weight on high-resolution features, consistent with the importance of lesion margins and local ultrasound texture. At the same time, the non-zero coarse contribution indicates that global tissue context remains useful. This balance provides a plausible mechanism for improving contour quality without discarding semantic information.

Boundary quality is particularly relevant to BI-RADS-oriented analysis because margin descriptors such as circumscribed, indistinct, angular, microlobulated, and spiculated depend on precise contour representation [23]. The 15.63-pixel HD95 obtained on the held-out test set therefore complements the Dice improvement by quantifying reduction in large boundary deviations. However, clinical interpretation of a pixel distance depends on acquisition geometry and image rescaling; prospective validation using physical spacing metadata is needed before translating pixel-level HD95 into millimetre-scale clinical claims.

The uncertainty analysis is promising but should be interpreted carefully. In the current implementation, entropy is derived from a spatial-attention signal and functions as an uncertainty proxy rather than a fully calibrated Bayesian predictive distribution. Its strong association with segmentation quality motivates further calibration experiments, comparison with Monte Carlo dropout or deep ensembles [13,14], and prospective testing of whether uncertainty-based case flagging improves radiologist workflow.

## 6. Limitations

- The main evaluation uses a relatively small public dataset (BUSI, n=647) from a limited acquisition setting; multi-centre external validation remains necessary.
- The five-fold experiment resamples the development set while keeping the same test set fixed. It measures training-set sensitivity but is not equivalent to a conventional five-fold cross-validation estimate over the entire dataset.
- The uncertainty signal is attention-derived and has not yet been demonstrated to be probabilistically calibrated.
- All baseline results are most comparable within the controlled training protocol used in this dissertation. Direct comparison with numbers reported in unrelated papers can be confounded by different splits, preprocessing, augmentation, and evaluation code.
- Runtime was measured on NVIDIA GPU hardware at 224×224 resolution; real-time clinical integration has not yet been prospectively validated on ultrasound systems or hospital workstations.
- The present study addresses binary lesion segmentation. End-to-end diagnosis, BI-RADS classification, multimodal fusion, and prospective clinical decision support remain future work.

## 7. Conclusion

This work presents UGB-SegNet, a compact breast ultrasound lesion segmentation framework combining multi-scale CBAM attention, learnable feature-pyramid fusion, an uncertainty-guided decoder, and boundary-aware deep supervision. On the BUSI held-out test set, the model achieved 81.30% Dice, 68.49% IoU, 81.71% sensitivity, 98.14% specificity, and 15.63-pixel HD95, while using 14.00M parameters and running at 62.4 FPS on the tested GPU. The controlled baseline comparison and component-level ablations indicate that the proposed design improves overlap, contour precision, and parameter efficiency. The next step is multi-centre external validation with calibrated uncertainty estimation and prospective assessment in clinical ultrasound workflows.

## Data Availability

All data used in the present study are publicly available. The study used the Breast Ultrasound Images (BUSI) dataset, an openly available de-identified breast ultrasound imaging dataset with corresponding segmentation masks. No new patient data were collected for this study.

https://www.kaggle.com/datasets/aryashah2k/breast-ultrasound-images-dataset

## Author Contributions

Taukir Alam: Conceptualization, Methodology, Software, Validation, Formal Analysis, Investigation, Data Curation, Visualization, Writing - Original Draft, Writing - Review & Editing.

Fang-Rong Hsu: Supervision, Methodology, Validation, Writing - Review & Editing.

## Code Availability

The implementation associated with this preprint is available at https://github.com/taukiralam007/UGB-SegNet. The repository can be updated alongside future arXiv versions to provide revised code, documentation, and reproducibility materials.

## Data Availability

BUSI is a publicly available dataset described by Al-Dhabyani et al. [4]. BUS-BRA is a publicly available breast ultrasound dataset described by Gómez-Flores et al. [19]. The CCH dataset discussed as Phase 1 background is not used for the primary UGB-SegNet evaluation in this preprint.

## Ethics Statement

The primary experiments in this preprint use publicly released, de-identified breast ultrasound datasets. The earlier Phase 1 CCH study referenced for research motivation was conducted under Changhua Christian Hospital IRB No. 181235, as reported in the dissertation and prior publication [24].

## Acknowledgments

The authors acknowledge the creators of the BUSI and BUS-BRA datasets and the open-source research community whose software and benchmark implementations supported this study.

## Notes

### Competing Interest Statement

The authors have declared no competing interest.

### Author Declarations

https://www.kaggle.com/datasets/aryashah2k/breast-ultrasound-images-dataset

